# Exploring the diversity of participants with dementia taking part in research: a mixed methods study

**DOI:** 10.1101/2023.09.06.23295133

**Authors:** Rupinder Kaur Bajwa, Louise Howe, Joy O. Agbonmwandolor, Alison Cowley, Emma J. Adams, Sarah Goldberg, Rowan H. Harwood

**Affiliations:** School of Health Sciences, University of Nottingham, Queen’s Medical Centre, Nottingham NG7 2HA2; School of Medicine, University of Nottingham, Queen’s Medical Centre, Nottingham NG7 2HA; Nottingham University Hospitals NHS Trust, David Evans Medical Research Centre, City Hospital, Nottingham NG5 1PB; School of Health and Social Care, University of Lincoln, Brayford Pool, Lincoln, LN6 7TS

## Abstract

**Introduction:** Dementia is becoming increasingly prevalent in the UK. Older adults from black and south Asian communities have a higher risk for dementia due to an increased prevalence of dementia specific risk factors such as hypertension, diabetes, and heart disease. Deprivation has also been linked to an increased risk of dementia. Ethnic minority and lower socioeconomic groups are underrepresented in dementia research. The aim of this study was to explore factors influencing diversity in dementia and rehabilitation research within the context of the Promoting Activity, Independence, and Stability in Early Dementia (PrAISED) randomised controlled trial (RCT).

**Methods:** We conducted an exploratory sequential mixed methods study to explore disparities in socioeconomic and ethnic diversity between the PrAISED RCT population and recruitment pathways used in one study site (Nottinghamshire) and compared these with regional and national data. We aimed to collate and summarise data available on ethnicity and deprivation for recruitment/referral pathways (Nottinghamshire site) and the PrAISED cohort (all sites). Additionally, we interviewed healthcare professionals (n=2), researchers (n=2) and members of black and south Asian communities (n=4) to explore barriers to participating in research for people with dementia.

**Results:** Under 2% of the overall PrAISED RCT sample (across all sites) were from a non-white ethnic minority background and a third of participants lived in areas with the least deprivation. Referrals to memory assessment services in Nottinghamshire included people from diverse socioeconomic backgrounds, with 7.3% being from non-white ethnic minority communities. Through interviews, several barriers to healthcare, research and rehabilitation were identified. Healthcare barriers included lack of awareness of dementia, mistrust, stigma, fear, and lack of culturally appropriate services. Research barriers included recruitment routes, awareness of research, language, and recruiter beliefs. Barriers to rehabilitation research included a lack of use of culturally appropriate language, more culturally specific barriers, and lack of representation.

**Conclusion:** Participants recruited to the PrAISED RCT were mainly white and socioeconomically privileged. Data recording and access around ethnicity is still inconsistent, making it difficult to ascertain at which point services and research become inaccessible for people from underserved communities. Future research needs to work with these communities to develop innovative solutions to overcome the barriers identified in this study and to put recommendations made into practice.

## 2 Background

Dementia is becoming increasingly prevalent within the UK population. In 2019, the Alzheimer’s Society reported an estimated 950000 people were living with dementia in the UK and this is set to double by 2040 (1). Dementia is a long term, degenerative neurological condition that will result in a loss of cognition, function and quality of life (2). A systematic review conducted by Public Health England (4), identified that dementia is more common in females, and in African-America, black-Caribbean or Hispanic groups. However, these groups are less likely to access assessment and diagnostic services in the same way as their non-minority ethnic peers (5). Similarly, Cooper et al (6) explored the uptake of anti-dementia medication and found that people from less deprived areas were 25% more likely to access medication as they were better able to negotiate health care systems. More recent work has highlighted a link between lower socioeconomic status (7) and increased risk of dementia, as well as poorer access to specialist diagnostic services (8).

It has been established that there is under representation of ethnic minority groups within healthcare research (29, 30) and for research into dementia care there is no exception (21). Two recent large RCTs of complex rehabilitation interventions for people with dementia reported only 4% (31) and 5-6% (28) of their overall participants were from ethnic minority backgrounds. There has also been under representation of people from a lower socio-economic background within healthcare research (32), although this is less thoroughly documented in dementia literature. The implication of having limited representation within a research trial is that the results will not necessarily be generalisable to sections of the population that were not robustly included within the study (33). There is a need to better understand how to increase diversity in research studies for individuals with dementia, to increase representativeness of the population and to ensure interventions meet the needs of diverse groups.

### 2.1 The PrAISED Trial

The Promoting Activity Independence and Stability in Early Dementia (PrAISED) trial was conducted between September 2018 and June 2022, to test a complex rehabilitation intervention for people newly diagnosed with mild dementia and mild cognitive impairment (3). Three hundred and sixty-five participants were recruited from five different geographical areas of England. Anecdotal information from the PrAISED therapists, researchers and PPI members identified that recruitment to the trial did not appear to be representative of the diversity that is known to be present across these sites.

### 2.2 Recruitment to the PrAISED trial

At the Nottinghamshire site, PrAISED participants were recruited from NHS Memory Assessment Services (MAS), the National Institute of Health Research’s Join Dementia Research (JDR) register, Clinical Research Networks (CRN) database and GP practices. Recruitment to the trial required participants to have a dementia or mild cognitive impairment diagnosis either through Memory Assessment services or other secondary healthcare services (Figure 1).

**Figure 1:**
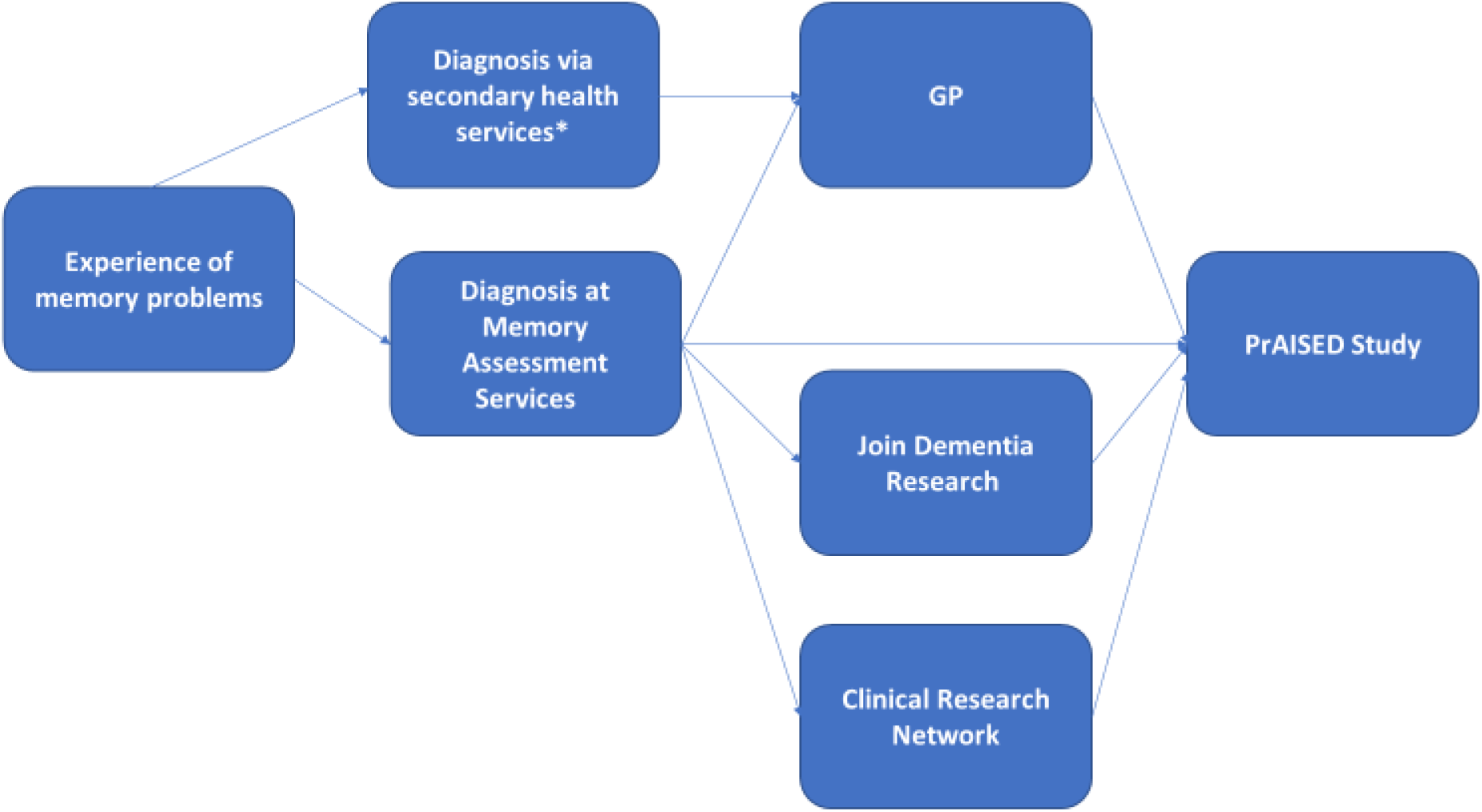
PrAISED Recruitment Pathway. **\***Diagnosis via secondary health services, i.e. Through community mental health team or acute general hospital admission.

This study aimed to explore factors influencing diversity in dementia and rehabilitation research within the context of the PrAISED RCT, using the Nottinghamshire study site as an exemplar case study.

## 3 Methodology

### 3.1 Study design

This study had an explanatory sequential mixed methods design, where the qualitative data were collected and utilised to expand on the quantitative findings (9). The quantitative data were collected as part of the RCT, and the quantitative data were analysed first to inform the interview schedules for the qualitative phase of the study. Following analysis, the qualitative findings were integrated with the quantitative findings to aid in interpreting the results (9).

### 3.2 Quantitative Methods

Data on ethnicity for the general population and the population aged 65 and over in England and Nottinghamshire was obtained from the Office for National Statistics Census 2021 (34). Data on the number of older adults aged 65+ with a diagnosis of dementia in England and Nottinghamshire was obtained from NHS digital (35).

For the PrAISED cohort, demographic data collected from participants enrolled in the PrAISED RCT, including gender, ethnicity, and years of education, were included in this study. Additionally, postcode data (where available) were used to identify area-level deprivation based on the English Indices of Multiple Deprivation (IMD) (10). The Index of Multiple Deprivation is a composite measure of seven domains: income, employment, education, health, crime, barriers to housing and services, and living environment. The IMD scores have been ranked using deciles with 1 being most deprived and 10 being least deprived.

Additionally, for the PrAISED recruitment pathways, data on ethnicity and IMD deciles were requested from MAS across Nottinghamshire, GP surgeries that took part in the PrAISED trial and the Join Dementia Research register (38)

For the MAS clinics recruitment pathway, the following data were collected for the PrAISED recruitment period (October 2018 to June 2021):

- Number, ethnicity and IMD decile for older adults aged 65+ referred to MAS.
- Number, ethnicity and IMD decile for individuals who attended and did not attend their initial appointment.
- Number, ethnicity and IMD decile for participants who received a diagnosis.

Data collected from the census, NHS digital, PrAISED cohort and PrAISED recruitment pathways were analysed using descriptive statistics in Microsoft Excel.

### 3.3 Qualitative Methods

Exploratory interviews were conducted with key stakeholders and community representatives. Key stakeholders were identified along the recruitment pathway, namely staff from MAS and the CRN who were the main facilitators to recruitment to the trial. The objective of interviewing stakeholders was to identify their experiences of working with people from diverse backgrounds who are living with dementia, accessing services, and participating in research. This study focused on the facilitators and barriers related to referral to service and recruitment to research studies.

Community representatives were also interviewed to gain their understanding of dementia, dementia services, rehabilitation, research and the barriers and facilitators to engaging in these areas. These interviewees were identified through existing community networks, public and patient involvement, and engagement links.

A pragmatic sample of two memory assessment nurses, two researchers from the CRN and four community representatives from different backgrounds was recruited. Semi-structured qualitative interviews were conducted between 30^th^September 2022 and 24^th^ October 2022. An example interview schedules is available in Supplement 1. The interviews were carried out in person or via the telephone and captured using a digital audio recorder by RB, JA and LH and lasted between 26 and 75 minutes. The recordings were transcribed verbatim. The interviews were uploaded to NVivo 12 (27) and analysed into codes and then themes using a five-step reflexive thematic analysis process (11). To ensure coding accuracy, 20% (n=3) of the interviews were coded by a second researcher and any discrepancies were discussed with the research team.

## 4 Results

### 4.1.1 Population statistics for England

18.4% (10,401,200) of the population in England are aged 65 and over (34). As of January 2022, 424,326 older adults aged 65 and over have a diagnosis code relating to dementia and it is estimated that the true number living with dementia is around 689,080 (35). Data shows 18% of people in England are from a black, Asian, mixed, or other ethnic group (34).

### 4.1.2 PrAISED cohort

The PrAISED RCT recruited 365 participant carer dyads from across 5 sites in England: Nottinghamshire, Derbyshire, Lincolnshire, Oxford, and Bath. 210 participants were male and participants on average had 12.5 years of education. Less than 2% (7/365) of participants were from a non-White ethnic minority community.

Three hundred and three participant carer dyads were recruited through MAS, 40 participant carer dyads were recruited through GP practices and 22 participant carer dyads were recruited through the JDR register. When looking at ethnic and socioeconomic diversity of participants by recruitment pathway, 0.3% (4/303) of participants recruited through MAS were from a non-white background, 4.5% (1/22) from the JDR recruitment pathway and 5% (2/40) for the GP recruitment pathway.

Over a quarter, 28% (104/365), of participants recruited to the PrAISED RCT lived in a neighbourhood that was within the 10^th^ (least deprived) decile of deprivation (, (Figure 2).

**Figure 2:**
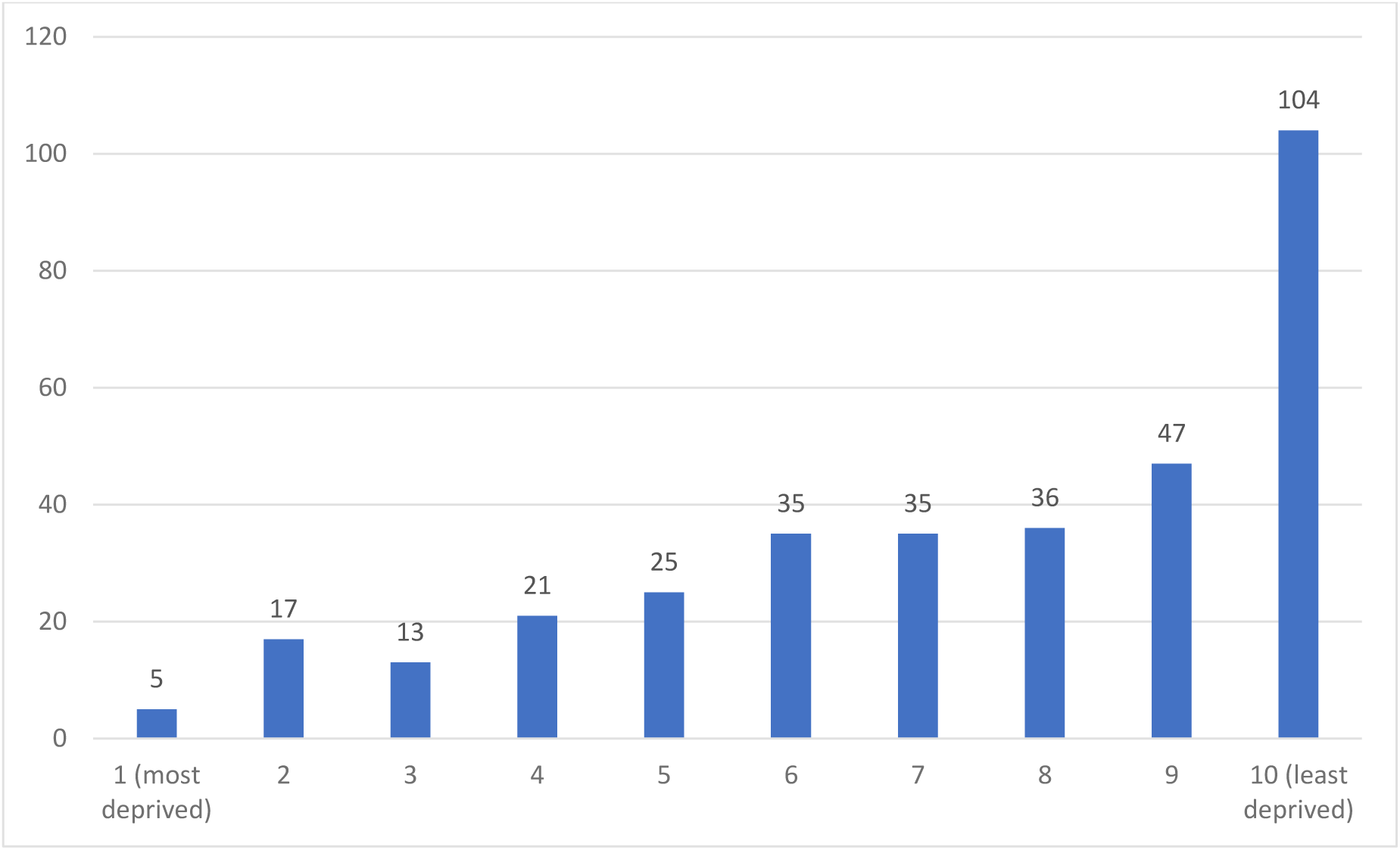
Count of IMD decile for PrAISED participants (all sites)

### 4.1.3 Nottinghamshire site as an exemplar case study

In Nottinghamshire there are 7,519 older adults aged 65 and over living with dementia, with the true figure estimated to be around 11,187 (36). Around 16% of the population In Nottinghamshire is from an ethnic minority community (34). Data from the 2021 Census for ethnicity of older adults aged 65 and over in Nottingham and Nottinghamshire has not yet been released by the Office for National Statistics.

#### 4.1.3.1 Memory assessment service recruitment pathway into the Nottinghamshire PrAISED site

Between October 2018 and June 2021, 4,910 referrals were made to MAS within Nottinghamshire were made. Of these referrals, 92.7% of individuals were white, 2% were Black (African, Caribbean, or Mixed White and Caribbean) and 2.5% were South Asian (Indian, Pakistani, Bangladeshi, or other, (Table 1). Eleven percent of referrals to MAS were from the most deprived post codes (556/4910) and 12% of referrals (632/4910) were from the least deprived post codes (Figure. 3).

**Figure 3:**
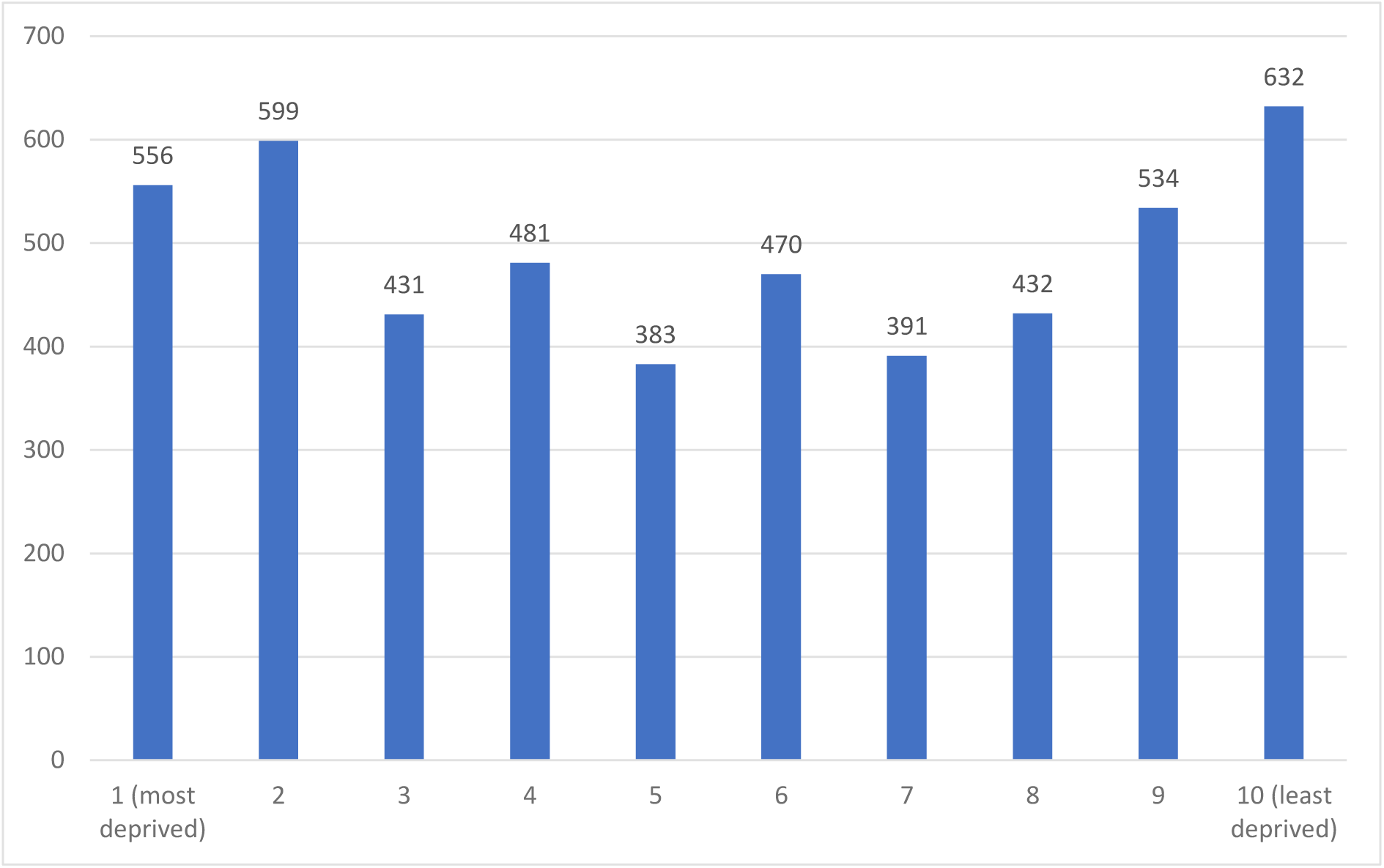
Count of IMD decile for MAS referrals between October 2018 and June 2021.

**Table 1.** Ethnicity of patients aged 65 and over referred to a memory assessment service during the PrAISED recruitment period (October 2018 to June 2021)

| **Referrals 65 and over** |  |  |
| --- | --- | --- |
| **Ethnicity** | **Count** | **%** |
| Arab | 1 | 0.0% |
| Asian/Asian Brit. – Bangladeshi | 2 | 0.0% |
| Asian/Asian Brit. – Indian | 52 | 1.1% |
| Asian/Asian Brit. – Other | 11 | 0.2% |
| Asian/Asian Brit. – Pakistani | 54 | 1.1% |
| Black/Black Brit. – African | 9 | 0.2% |
| Black/Black Brit. – Caribbean | 76 | 1.5% |
| Black/Black Brit. – Other | 10 | 0.2% |
| Chinese | 8 | 0.2% |
| Mixed – Other | 27 | 0.5% |
| Mixed – White & Asian | 2 | 5.1% |
| Mixed – White & Black Caribbean | 6 | 0.1% |
| Not Known | 38 | 0.8% |
| Not Stated | 39 | 0.8% |
| Other Ethnic Group | 21 | 0.4% |
| Romany Gypsy | 1 | 0.0% |
| White – British | 4447 | 90.6% |
| White – Irish | 45 | 0.9% |
| White – Other | 61 | 1.2% |
| Grand Total | 4910 |  |

Of those referred to MAS, 3,567 individuals attended their appointments, of whom 92.7% identified as White, 2.2% identified as Black and 2.8% identified as South Asian (see table 2). Four hundred and eleven individuals (11.5%) who attended referral appointments lived in postcodes in the 1^st^ decile of deprivation and 466 individuals (12.7%) who attended referral appointments attended were by individuals lived in areas in the 10^th^ decile of deprivation (figure 4).

**Figure 4:**
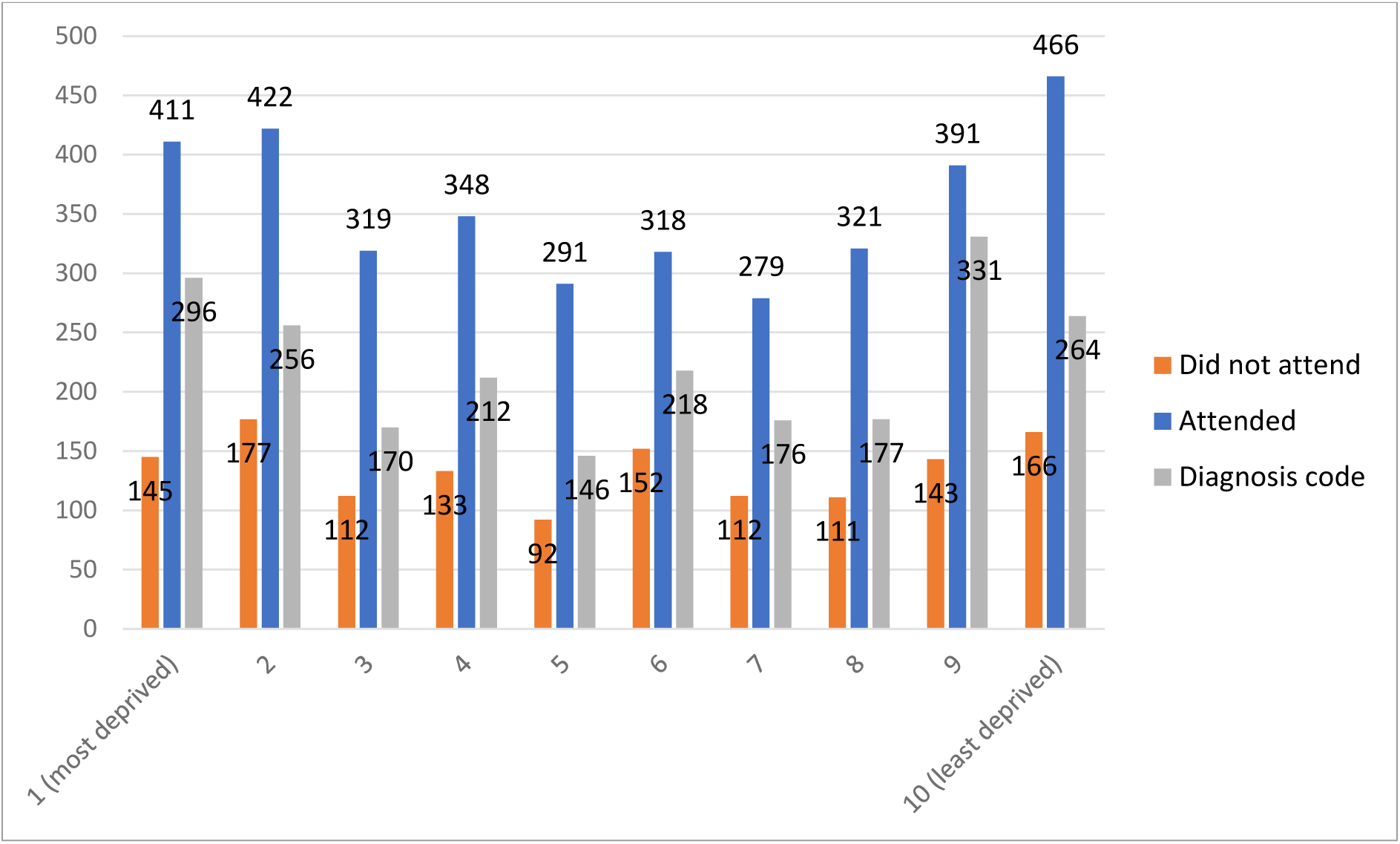
Count of IMD deciles for patients referred to MAS who did not attend the initial appointment, patients referred who attended the initial appointment, patients referred for whom a dementia diagnosis code was recorded.

**Table 2:**
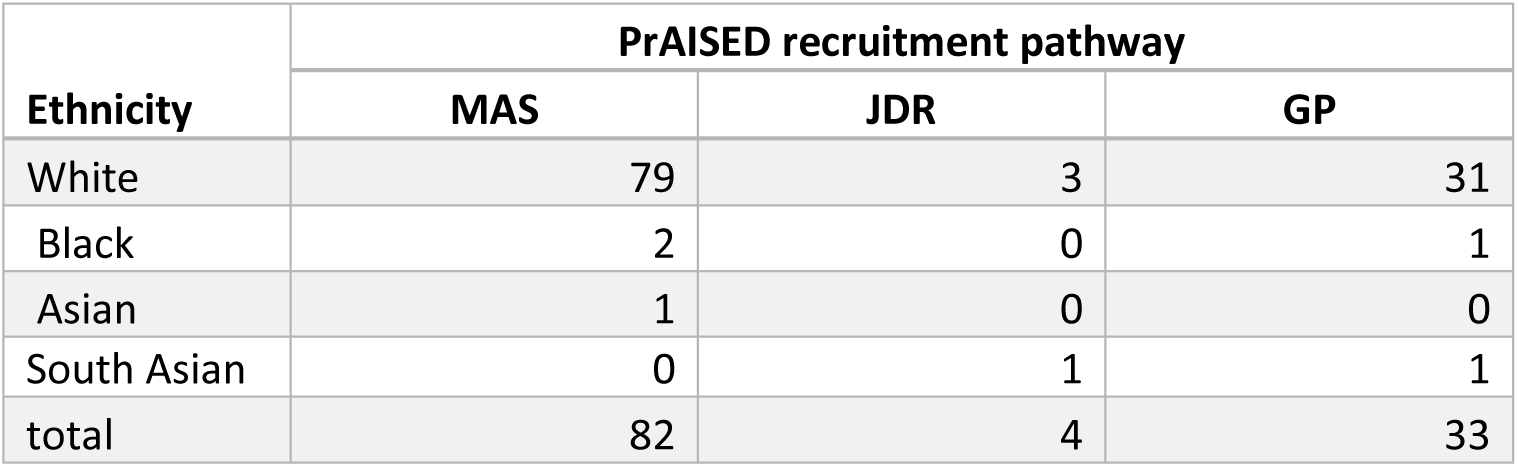
Breakdown of participant ethnicity based on recruitment pathway for Nottinghamshire site.

| Ethnicity | PrAISED recruitment pathway |  |  |
| --- | --- | --- | --- |
|  | MAS | JDR | GP |
| White | 79 | 3 | 31 |
| Black | 2 | 0 | 1 |
| Asian | 1 | 0 | 0 |
| South Asian | 0 | 1 | 1 |
| total | 82 | 4 | 33 |

Of the 1,343 individuals who were referred but who did not attend their appointments, 92.7% were White, 1.5% were Black and 1.4% were South Asian (see table 2). Of those appointments that were not attended,145 referral appointments not attended were from individuals that lived in areas in the 1^st^ decile of deprivation and 166 referral appointments not attended were from individuals that lived in areas in the 10^th^ decile of deprivation (figure 4).

Between October 2018 and June 2021, 2250 referrals had a diagnosis code relating to dementia, of these 95.8% were White, 1.4% were Black and 0.6% were South Asian (see table 2). There were 296 referrals with a dementia diagnosis code from the 1st decile for deprivation and 264 referrals with a dementia diagnosis code were from the 10th decile for deprivation (Figure. 4).

**Table 1.** Ethnicity of individuals referred to MAS for patients referred to MAS who did not attend the initial appointment, patients referred who attended the initial appointment, patients referred for whom a dementia diagnosis code was recorded.

| **Ethnicity** | **Attended** | **%**<br>**Attended** | **DNA** | **%**<br>**DNA** | **Diagnosis**<br>**code** | **%**<br>**Diagnosis**<br>**code** |
| --- | --- | --- | --- | --- | --- | --- |
| Arab | - | - | - | - | 1 | 0.0% |
| Asian/Asian Brit. - Bangladeshi | 2 | 0.1% | - | - | 1 | 0.0% |
| Asian/Asian Brit. - Indian | 43 | 1.2% | 9 | 0.7% | 9 | 0.4% |
| Asian/Asian Brit. - Other | 10 | 0.3% | 1 | 0.1% | 1 | 0.0% |
| Asian/Asian Brit. - Pakistani | 45 | 1.3% | 9 | 0.7% | 4 | 0.2% |
| Black/Black Brit. - African | 5 | 0.1% | 4 | 0.3% | 4 | 0.2% |
| Black/Black Brit. - Caribbean | 62 | 1.7% | 14 | 1.0% | 26 | 1.2% |
| Black/Black Brit. - Other | 8 | 0.2% | 2 | 0.1% | 2 | 0.1% |
| Chinese | 6 | 0.2% | 2 | 0.1% | 1 | 0.0% |
| Mixed - Other | 21 | 0.6% | 6 | 0.4% | - | - |
| Mixed - White & Asian | 2 | 0.1% | - | - | - | - |
| Mixed - White & Black African | - | - | - | - | 1 | 0.0% |
| Mixed - White & Black Caribbean | 5 | 0.1% | 1 | 0.1% | 1 | 0.0% |
| Not Known | 17 | 0.5% | 21 | 1.6% | 15 | 0.7% |
| Not Stated | 17 | 0.5% | 22 | 1.6% | 11 | 0.5% |
| Other Ethnic Group | 15 | 0.4% | 6 | 0.4% | 15 | 0.7% |
| Romany Gypsy | 1 | 0.0% | - | - | - | - |
| White - British | 3235 | 90.7% | 1212 | 90.3% | 2089 | 92.8% |
| White - Irish | 34 | 1.0% | 11 | 0.8% | 23 | 1.0% |
| White - Other | 39 | 1.1% | 22 | 1.6% | 46 | 2.0% |
| **Total** | **3567** |  | **1342** |  | **2250** |  |

#### 4.1.3.2 GP recruitment pathway in Nottinghamshire

In Nottinghamshire, 114 GP practices (41 in Nottingham city and 73 in Nottinghamshire County) were approached about participating in a search, screen, and mail out for the PrAISED RCT. Of those approached, 32 practices participated (13 in Nottingham city and 19 in Nottinghamshire County) and a total of 880 patients were mailed further information about the PrAISED RCT. For this study, we re-contacted a selection of GP practices that contributed to the PrAISED recruitment to collect data on ethnicity, however, were not able to collect this data as practice managers advised it is not routinely collected by practices during the patient registration process.

#### 4.1.3.3 Join Dementia Research pathway in Nottinghamshire

We were unable to access data on the ethnic and socioeconomic diversity of volunteers with memory problems who were registered on the JDR website during the PrAISED recruitment period.

A recent report by the Alzheimer’s Society found that those from ethnic minority communities were under-represented on JDR (38).

#### 4.1.3.4 PrAISED Nottinghamshire site

The Nottinghamshire site recruited 119 participant carer dyads. Overall, 113 participants identified as White, 3 identified as Black, 3 Identified as South Asian. Of these participants, 82 dyads were recruited from MAS clinics across Nottinghamshire (excluding Bassetlaw), 4 dyads recruited from JDR, and 33 dyads recruited through GP practices. Table 2 shows the breakdown of participant ethnicity for each recruitment pathway. When looking at where participants were recruited from in terms of socioeconomic deprivation within Nottinghamshire, 42% (50/119) of participants recruited in Nottinghamshire were from an area that was within the 10^th^ IMD decile (Figure. 5).

**Figure 5:**
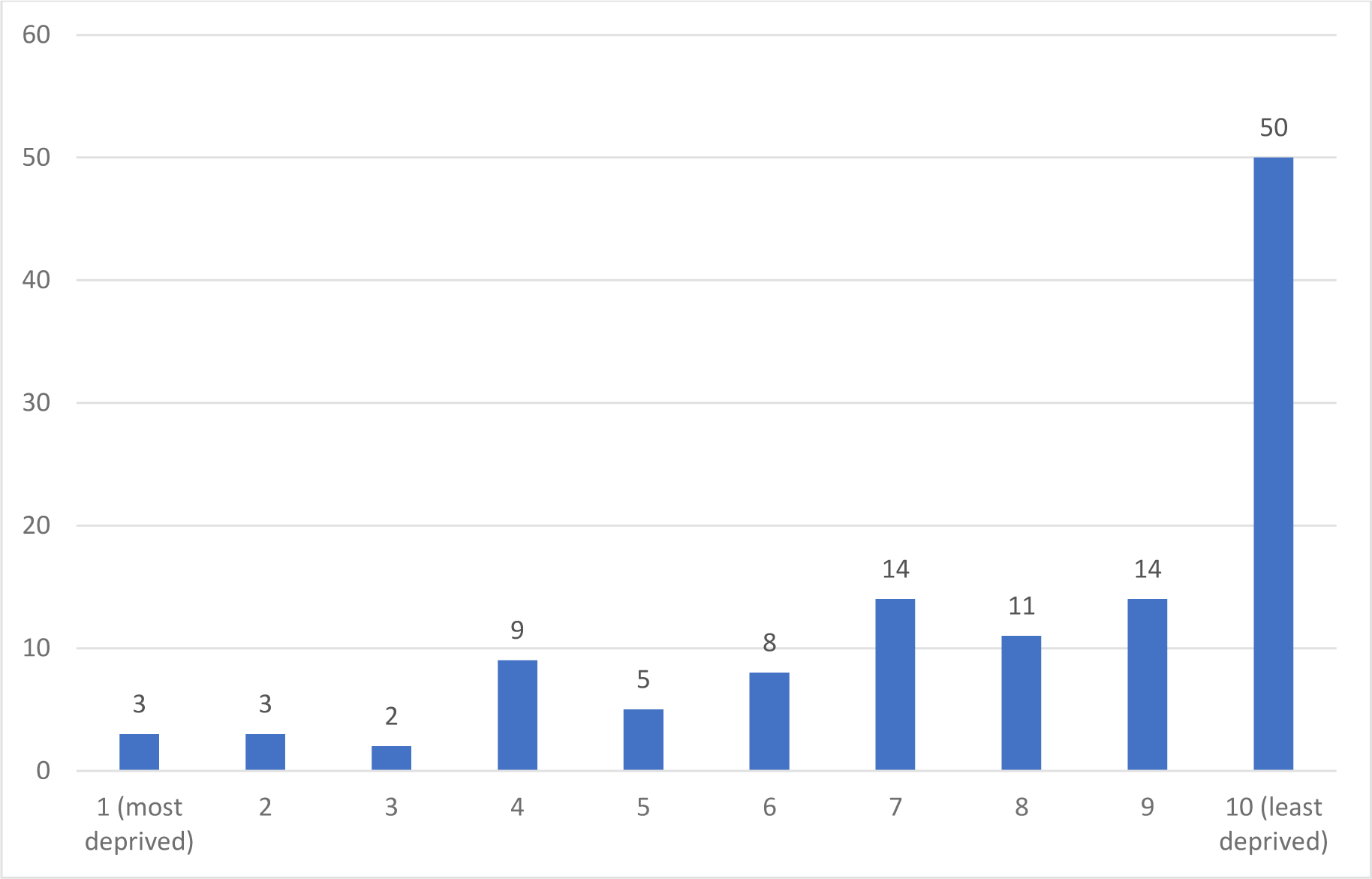
Count of IMD decile for PrAISED participants (Nottinghamshire site)

### 4.2 Qualitative Findings

Interviews were completed with eight interviewees who were allocated participant numbers P1 through to P8. Table 3 shows which organisations or communities participants were recruited from.

**Table 3:** Qualitative Interview Participant Demographics.

| Representation | Number of interviewees |
| --- | --- |
| MAS | 2 |
| CRN | 2 |
| Jamaican Community | 1 |
| South Asian Muslim Community | 1 |
| Pakistani Community | 1 |
| Sikh Community | 1 |

The qualitative findings have been broken down into two broad themes, awareness and beliefs about dementia and services, and a lack of cultural awareness and consideration. Associated with these themes are sub themes (table 4).

**Table 4:** Themes and Sub Themes derived from the qualitative interviews.

| Theme | Subtheme |
| --- | --- |
| Awareness and beliefs about dementia and services | Awareness of dementia |
|  | Trust in healthcare professionals and the system, stigma, fear |
| Lack of cultural awareness and consideration | Lack of culturally aware services<br>• Specific cultural barriers<br>• Representation<br>• Language |
|  | Access to Services |
|  | Research Methods<br>• Recruitment routes<br>• Lack of awareness of research<br>• Referrer/Recruiter's beliefs |

#### 4.2.1 Awareness and beliefs about dementia and services

different communities had a different understanding about dementia as a condition and different perceptions about healthcare services.

##### 4.2.1.1 Awareness of dementia

Many of the interviewees described differing levels of awareness of dementia in minority ethnic communities. For example, one interviewee described his community as seeing dementia as a ‘Westernised condition’ due to different attitudes to caring for older people.

> *‘And we’ve had discussions about dementia and a lot of people I think in the Muslim community, at least in Nottingham, talking to Muslims, they can think it’s a westernised condition. It’s a white person’s condition because they don’t look after their elderly the way they do. OK. And it’s often seen that way, rather than a cognitive functioning or, you know, a neurological impairment or like you have vascular dementia, lack of oxygen, all those things, instead of looking for the condition, they’re looking at it through a different lens’ (I7)*.

Cultural differences were considered by another interviewee as ‘masking’ the condition because the person with dementia lived with other generations who compensated for their declining abilities. It was also thought that those from an older minority ethnic generation put cognitive difficulties in older age down to their age and lack of education, rather than a potential illness. Having different understandings or beliefs about old age and dementia meant that for some groups of people accessing healthcare or obtaining a diagnosis of dementia was not pursued.

##### 4.2.1.2 Trust, stigma, and fear

Interviewees highlighted the difficulties of some communities trusting health care services. One research representative described how people from lower socio -economic backgrounds can begin to feel suspicious of institutions from an early age and that this can result in a distrust of the NHS and other services.

> *‘The working class or the people on the poverty line have lost that trust. Some of them it’s instilled in them very early on, right from when they’re this high, so when they’re four, five we don’t trust the police, we don’t trust this, we don’t trust that we don’t trust this. We don’t trust the NHS’ (I7)*.

There was also some reticence to access healthcare services for people with dementia due to a fear of diagnosis and the stigma this might bring. For example, one interviewee discussed how his family had been in denial regarding a family member’s diagnosis and that this was kept hidden from the local community. It was felt this may have a wider implication for the family including marriage prospects for younger members.

> *‘So I think there needs to be, but that’s the negative side of it isn’t it really when you think about it, oh God this person’s unwell, they’ve got an illness, which means we should keep away from them, our children must keep away from there. How are our children or our grandchildren’s prospects of marriage knowing that when they turn old they could have dementia which means that my daughter or my son will have to take care of that person, we don’t want the burden of our children going through looking after their partner in that way. So I think there needs to be a little bit more exploration into language which is not taboo’ (I3)*.

#### 4.2.2 Lack of cultural awareness and consideration

The second broad theme within the qualitative data was that of a lack of cultural awareness and consideration, this could be broken down into further subthemes including specific cultural barriers, a lack of representation, language barriers, difficulties accessing service and unaccommodating research methods.

Interviewees felt that there was at times a disconnect between cultural needs and service provision. A language barrier was strongly acknowledged, this resulted in problems with diagnosis, significant miscommunications, and a reliance on family to interpret. There was a general feeling from community representatives that they were not listened to and that healthcare professionals felt they were too difficult to deal with. Two interviewees identified these as reasons for an excessive length of time between when concerns were first highlighted to the GP and their family members becoming diagnosed with dementia.

> *‘And I think the kind of, when you have things like a language barrier, then what ends up happening is that people just decide well you’re a bit too difficult to kind of deal with so we’re not going to deal with you so just be grateful for what it is that you actually get in the way of treatment if any’ (I3)*.

##### 4.2.2.1 Lack of culturally aware services

###### Specific cultural barriers

In certain healthcare situations, there were other barriers for example the need to be segregated by sex, that were not always met. Interviewees described how older people did not feel fully accepted in healthcare settings because of their culture and religion, especially if there were language barriers.

Interviewees also identified specific cultural barriers that may affect members of different cultural groups engaging with rehabilitation or exercise. These would include cultural and religious practices around clothing, jewellery and restrictions on how and when to undress. There may also be concerns or restrictions that mean exercise as a group, particularly that of mixed gender participants may not be acceptable. This may also include the gender of any professionals supporting the individual with the intervention.

> *‘I think that would work a lot better, but then I think that it should also be designed where it’s a same sex individual. So the person coming to deliver the programme is of the same sex as that person. Just because obviously biologically different but also the comfortability of having a stranger in one’s own kind of environment, it makes it a little bit easier’ (I3)*.

###### Representation

The issue of representation was also highlighted through the interview. Often research and healthcare staff are from a different background to the individual, thus the participant may feel like these services are not something that is relevant to them. This added to the believe that they won’t be understood and accepted for who they are.

*I mean the things we can’t control, I suppose if you had researchers or clinicians that were from the same community or same background as the people that you’re wanting to participate, that would help, but that’s something we don’t really have any control over (I4)*.

###### Language

The difficulties with language were challenging especially if they were accompanied with a lack of literacy skills. This combined with a lack of using culturally acceptable terms were identified as reasons why health education may not be accepted. One community member identified that there was-

> *‘a massive educational barrier between the lived experience of my aunt who doesn’t speak English as her first language and is also a practising Muslim versus the kind of white British way of providing services for somebody that doesn’t come from the same background ‘ (I3)*

Language is a substantial barrier for people participating in research. Being able to communicate in English was a requirement of the PrAISED trial and one interviewee felt that this may have put people off if they were less confident in their language skills. There may also be difficulties relating to literacy skills which can be problematic in a research context where a lot of written information is involved. Another interviewee identified that the use of REC (Research Ethics Committee) approved documents which are typically lengthy and technical may exclude people from different backgrounds.

> *‘Who are the REC, who are they? I’ll tell you who they are. They’re predominantly white middle class people, OK, and they approve our documentation which we then send out into the communities to try and be inclusive. OK, it’s dry. It’s too dry and it’s excluding people from the studies, because we’ve only got this approved document which we can use, and we then go out into the communities and we’re not really recruiting as effectively as we should be’ (I7)*.

Alongside language difficulties are that of a lack of education and literacy skills. This means that translating written materials may not make them accessible to the potential recruit.

> *‘I suppose, I know people have gone for things like making sure the resources are translated, but again, and you could do some, but I’m not sure how many people would be able to, especially in our Pakistani community, there isn’t that many people that could read Urdu, which is the language that they read. So that’s an issue and it might mean that they might not be able to read it’ (I3)*.

The current use of technical, formal language and language requirements in trials are other factors the interviewees felt were excluding groups of potential recruits to research trials. The interviewees highlighted the need to use language that is culturally acceptable. For example, one interviewee thought that the use of the term ‘exercise’ may have negative connotations to elderly members of their community who don’t see themselves as engaging in exercise, even if they did participate in active pursuits such as walking and swimming.

##### 4.2.2.2 Access to services

Those of lower socio-economic status were thought by interviewees to be less likely to attend MAS.

People referred to MAS are required to attend multiple appointments which are not always in the same place, or on a bus route. Therefore, people who didn’t have access to transport or taxis had more difficulty attending appointments.

It was also identified that most people who attended the MAS clinics, did so with the support of a family member or carer. People with spouses, and who were retired, were more likely to have family assistance in making and travelling to appointments than those that did not live with family members. People who were supported by younger family members who were also in employment and supporting children had less time to support their loved ones with appointments.

> *‘People from different ethnic backgrounds. Maybe there’s more, I’m thinking more sons and daughters presenting with mothers and fathers, who again would be interested in that study, but perhaps haven’t got the same time commitment to be there. Coming to an appointment with us is sometimes difficult for people, coming to a follow-up is difficult for people to take time out’ (I2)*.

##### 4.2.2.3 Research methods

###### Recruitment routes

Several interviewees identified that typical recruitment routes based on the medical model were not facilitating access for a wide range of people and therefore people from minority ethnic communities or a lower socio-economic background were not being exposed to recruitment materials.

*They’re {Researchers] not going through networks that are necessarily representative of society, what they’re doing is going through what they already know, and these are already known pathways that are predominantly white British’* (I7).

Problems with recruitment were also discussed in terms of the inclusivity of recruitment materials. It was generally felt that not everybody would be included if research required participants to go online, due to a lack of skills or a lack of available technology.

Interviewees highlighted the need for recruitment to include incentives for participants. This could be a financial reimbursement for those who need to take time out from work or incur costs of travelling, to be able to participate in a research study. As well as financial incentives, participants highlighted that it needed to be clear what the individual benefits for taking part in the research are.

> *‘Yeah and I suppose having an animation, making a video or if you wanted people from that community in the video in their language, maybe talking about the study and what it is and why it’s beneficial. Because then people could see that as well, but they can also hear it’ (I5)*.

###### Lack of awareness of research

The interviews highlighted that many people have misconceptions about what research involves, for example that this would usually involve medication or injections, and this can make people fearful and reluctant to take part. One community representative identified that she had never been invited to participate in research and another felt that if he had a lack of knowledge about research, those who accessed fewer services and were less articulate or had difficulties with the English language were even less likely to be aware of what research they could be involved in.

###### Referrer/Recruiter’s beliefs

Finally, another potential barrier that was discussed in the interviews was that of the referrer or the recruiter’s beliefs. One interviewee referred a lot of people to the PrAISED trial however acknowledged that other colleagues across the county may not have been enthusiastic about it when presenting it to potential recruits. This may have affected uptake of the trial in certain areas of the county. It was also identified that referrers may be gatekeeping which individuals they give information to. Another interviewee described clinicians as wanting to protect their patients who they feel have been experiencing challenging situations by not providing them with information about research that they may be eligible for. In this way potential research participant cohorts may be influenced by biases prior to recruitment.

> *‘What’s the issue is they’re providing this protective, in clinical practice, providing this, what I think is a protective cloak if you like over these people with dementia. And I’m not even going to approach them and ask them about this because I don’t think it’s relevant because they’ve gone through enough. Even though they’ve got capacity and they’re not approaching them, even though it’s denying them the right to be part of research if they want to be…’ (I7)*.

## 5 Discussion

This study aimed to explore factors influencing diversity in dementia and rehabilitation research within the context of the PrAISED RCT, using the Nottinghamshire study site as an exemplar case study using a mixed methods approach.

The findings from this study identified that older adults living with dementia from ethnic minority groups and in areas of deprivation were underrepresented in the PrAISED RCT. In the trial under 2% (7/365) of the population were from a non-white ethnic minority group compared to 18% in the general population in England. Additionally, nearly a third of participants (104/365) were living in a neighbourhood in England that was within the 10^th^ IMD decile (the least deprived areas in England). For the Nottinghamshire site specifically, although the inclusion of participants from ethnic minority communities demonstrated better representation (6/119), nearly half of the participants recruited within Nottinghamshire (50/119) were from an area within the 10^th^ IMD decile (least deprived areas in England) and there was a lack of socio-economic variation.

From the qualitative analysis, the identified communities had many barriers to accessing research. Firstly, due to a reliance on using healthcare as a route to recruitment, a lack of engagement in healthcare services had similar repercussions for research. There was a distrust of healthcare services, concerns about the stigma of a dementia diagnosis and a lack of culturally sensitivity within some healthcare services that may be leading to a lack of engagement in Memory Assessment Services. Restrictions to recruitment based on language also purported to be excluding otherwise suitable candidates which would widen the diversity within recruitment samples. Accessing rehabilitation research had further barriers related to feelings of a lack of representation and interventions that accommodate cultural requirements, particularly for South Asian communities.

The limited inclusion of people from black and South Asian minority groups and diverse socioeconomic background is not unique to the PrAISED RCT, with studies such as the DAPA trial and FINALEX also recruiting largely white populations (12,13, 28, 31). Systematic reviews exploring representation of ethnic groups in dementia trials also found that non-white ethnic minority groups were underrepresented (14,15) with one review reporting on average 94.7% of study populations in drug trials for Alzheimer’s disease between 2001 and 2019 were White. Underrepresentation of diverse groups is not unique to dementia research, with other areas of medical research highlighting the historic underrepresentation and need for increasing access to research for minority groups (16–18).

When exploring ethnic and socioeconomic diversity within the PrAISED recruitment pathways at the Nottinghamshire site, we were only able to access data on ethnicity and deprivation from the MAS pathway. GP surgeries involved in screening for PrAISED recruitment did not routinely collect ethnicity data from patients during the registration process. We also were unable to access data on ethnicity and socioeconomic status volunteers registered on JDR in Nottinghamshire. Though a recent report by the Alzheimer’s society (19)and a study from JDR (39) have identified that areas of deprivation and minority communities are underrepresented on the JDR. These findings highlight a bigger challenge around data recording and access. Previous work has also highlighted that ethnicity is not well recorded in research and highlights the need for improving data capture and reporting on ethnicity (14, 20, 42). Without routinely recording this data to enable monitoring and uptake of dementia services and participation in research by marginalised groups, it is a challenge to ascertain which groups are being excluded, at what stage and where more targeted engagement strategies are required.

This study echoed similar findings into the experience of minority ethnic communities accessing healthcare and research for people with dementia. A meta-analysis of 33 studies identified that within Western society those from a minority ethnic background are diagnosed at a later stage and less likely to access healthcare and participate in research (43). The barriers identified to recruitment above are similar to those previously reported in a systematic review, which also highlighted barriers related to health services, research processes as well as practical and community related barriers (21, 41). Differences between cultural understanding of dementia, shame, stigma and negative experiences of healthcare services have previously been identified as barriers to people from minority ethnic communities accessing diagnostic services (22, 44, 45). Previous work has also discussed the issues of stigma relating to dementia for ethnic minority groups, especially within south Asian communities and the impact of this not only for the individual but for their family unit (21,22)

In addition to the barriers discussed above, this study also found additional barriers around lack of trust and culturally aware services. Our interviewees also highlighted the lack of culturally inclusive healthcare organisations and the need for culturally aware staff to be able to cater for the needs of diverse communities, which has also been highlighted by a previous meta-synthesis of qualitative studies exploring barriers to dementia care for ethnic minority groups. (23). The issues around trust of healthcare professionals and services are not unique to dementia services. Trust can be difficult to build in intercultural healthcare and research practices due to historic cases of unethical research practices in the black community (50), personal negative experiences of healthcare (40, 22), and a fragmented, discontinuous model of healthcare delivery.

This study also highlighted research specific barriers to participation including lack of awareness of what research is, what it involves and what the benefits are for the individuals. The requirement to speak English to participate and the style of language used in research documentation (such as information and consent forms) can be limiting for older adults from minority communities where literacy skills may be poor or where English may not be their spoken language. Additionally, for those who do speak English, research language may involve technical terms or jargon which may put people off from taking part. Research methods and processes may also hinder inclusion as current systems for study document approvals are rigid and time consuming. Recruitment routes used in research such as recruiting from healthcare services is another barrier. Previous work has shown that older adults with memory problems from ethnic minority and lower socioeconomic groups do not access healthcare services until later in the disease progression, or may only access services when in crisis, at which point they may not meet strict eligibility criteria (22,24). A qualitative systematic review exploring recruitment and methodological issues in dementia research in ethnic minority communities reported similar themes around the use of language and recruitment strategies being barriers to research participation (21).

Another noteworthy subtheme relating to research barriers was the recruiter’s beliefs and their priorities. If the recruiter or healthcare professional were not enthusiastic about the study, this may have deterred patients. Additionally, recruiters and healthcare professionals may act as gatekeepers prioritising protection of patients they may perceive as in a state of hight stress therefore limiting their access to recruitment materials. The National Standards for Public Involvement (47) identify that we should be offering inclusive opportunities for people to engage in research.

To date there has been little exploration around the barriers to rehabilitation research for older adults with memory problems from ethnic minority communities and diverse socioeconomic groups. We found several barriers to rehabilitation research around the lack of culturally appropriate language, consideration for specific cultural barriers such as traditional dress which could impact an individual’s ability to take part in particular rehabilitation activities and the lack of representation of staff delivering rehabilitation programmes. These findings are in line with previous work which has highlighted cultural barriers to physical activity and exercise participation. Two systematic reviews of barriers and facilitators to physical activity in ethnic minority groups in the UK found that religion may facilitate participation in physical activity, but religious fatalism may be a barrier (51,52). Mixed gender exercise classes and dressing in exercise type clothing may also not be socially acceptable in some cultures. The lack of culturally aware spaces for physical activity was another key barrier to participation (51,52). Researchers clinicians and funders should look to address these barriers when designing new rehabilitation interventions to ensure that future rehabilitation research is accessible and inclusive.

### 5.1 Limitations

Previous research has shown that people from ethnic minority and lower socioeconomic background often access service at a later stage or crisis point and often experiences challenges and delays during the diagnosis process (5,24–26). We did not collect data from memory services on the dementia severity of patients who received a diagnosis. Including this data would be useful to understand at what stage of dementia people from minority groups are presenting in the health service and could explain why these groups may be underrepresented in the PrAISED RCT which aimed to recruit older adults with mild cognitive impairment and mild dementia.

Challenges around data recording and access meant we could not fully explore diversity present in all recruitment pathways for PrAISED in Nottinghamshire, thus making it difficult to ascertain where services and research become inaccessible for ethnic minority groups and those from diverse socioeconomic backgrounds. GP practices reported that they did not collect data related to ethnic group meaning we were not able to assess this data. Future work needs to look at how data capture of diverse characteristics can be improved to ensure that research and services in dementia and wider can monitor access and uptake of healthcare services and research and improve engagement with underserved communities at a local and national level.

The interview sample size was small. We spoke with three interviewees from the South Asian community and one participant from the Black Caribbean community. It is important to note that there are lots of different communities within Black and South Asian groups and though there are common barriers experienced across communities, there may also be unique community-specific barriers which we did not identify. These interviewees were identified through existing Patient and Public Involvement networks and therefore already had some knowledge of research processes, this may have added to their understanding of the topic, however their opinions may be different to members of their community with less understanding. Future work should look to examine unique challenges for each community in more detail.

The research interviewees representing MAS for example were based within areas of the county where there was less diversity and had received a positive reception to discussing research participation to people with dementia. Attempts were made to recruit representative of more deprived areas of Nottinghamshire, however there were no established connections with these communities. It was identified that recruitment would have required the researchers to have built community relationships and developed trusting partnerships with community organisations to develop a recruitment pathway and this was not possible within the time frame of the study.

A series of recommendations for dementia research and clinical research more generally are outlined below to improve the diversity of ethnic minorities and lower socioeconomic groups in research. Improving equity, diversity, and inclusion of older adults with memory problems from historically marginalised communities in dementia services and research is important to ensure that all those affected by dementia can live well with dementia.

### 5.2 Recommendations

Based on the findings, some recommendations are proposed for services and research both for people with dementia and the wider population

- **Increase representation and develop a culturally competent workforce** (33, 49). Funders and Universities should invest in developing opportunities for diversity in their workforce to enable research teams to look similar to the target participants and speak their languages. Developing workforce skills in cultural competence will enable participants to feel understood and assured that their needs will be met.
- **Build trust between communities**. Investment is required from universities and healthcare services to provide stable resources and staff to build these relationships. Mistrust in minority communities is a key barrier to service access. To build trust, healthcare professionals and researchers need to adopt a non-judgemental approach, awareness, and acceptance of other’s cultures (33, 46, 48, 49). Additionally, improving awareness of dementia, related services and research available to these groups will also aid inclusion and access.
- **Provide accessible information** such as videos with study information in addition to written participant information sheets. Information sheets and consent forms need to be shorter and simpler. This may be helpful for groups where English may not be their first language, or for groups who may have a lower level of literacy (33, 46, 48). Translate information into different languages for those who do not speak English and ensure the style of language used is in line with the style used by different groups in their day-to-day interactions (33). It is important to ensure that the information presented uses culturally acceptable terms.
- **Develop recruitment strategies**. These need to be flexible, rather than just clinic or service-based, researchers should go out into the community and settings that are regularly attended by ethnic minority groups (33, 47, 48). For instance, the radio may be an effective strategy to advertise research and disseminate findings to older adults from Black and South Asian communities. Targeted promotion may be a useful strategy to increase inclusion and access for certain groups that may be consistently underrepresented.
- **Provide renumeration**, particularly for lower socio-economic groups (46). This may improve the accessibility and subsequently inclusion of these groups in research. Potential participants from lower socio-economic groups may face the dilemma of lost earnings and being out of pocket if they take the time to participate in research, covering expenses and lost earnings can improve access for these groups.
- **Increase flexibility within research methods and processes.** Current process for approvals for studies and related documents are often rigid and can take a long time. Working with research governance bodies and including stakeholders from multiple communities a more dynamic research processes need to be developed so that research can be adapted quickly to meet the needs of different communities (46, 48).

## 6 Conclusions

This study has highlighted the disparity between diversity in the community, in referrals to services and in diversity in research studies, drawing on the PrAISED research programme as an example.

Gaps in data recording and access to obtain information about ethnicity and deprivation have been noted. Several barriers were identified at different points in the healthcare and research systems for Black and South Asian ethnic minority groups. Researchers need to work with ethnic minority and socioeconomic diverse groups to explore common and community-specific barriers to access and inclusion in dementia services and research. In addition to this, future work should include working closely with underserved communities to develop and implement actions to address barriers.

### Statement of ethical approval

Ethical Approval: This study was approved by the Bradford-Leeds Research Ethics Committee (REC number 18/YH/0059, IRAS project identification 236099) and a substantial amendment was gained to enable the collection of further historical quantitative data and qualitative interviews.

## Data Availability

All data produced in the present study are available upon reasonable request to the authors

